# Infectious viral load in unvaccinated and vaccinated patients infected with SARS-CoV-2 WT, Delta and Omicron

**DOI:** 10.1101/2022.01.10.22269010

**Authors:** Olha Puhach, Kenneth Adea, Nicolas Hulo, Pascale Sattonnet, Camille Genecand, Anne Iten, Frédérique Jacquérioz Bausch, Laurent Kaiser, Pauline Vetter, Isabella Eckerle, Benjamin Meyer

## Abstract

**Background:** Viral load (VL) is one determinant of secondary transmission of SARS-CoV-2. Emergence of variants of concerns (VOC) Alpha and Delta was ascribed, at least partly, to higher VL. Furthermore, with parts of the population vaccinated, knowledge on VL in vaccine-breakthrough infections is crucial. As RNA VL is only a weak proxy for infectiousness, studies on infectious virus presence by cell culture isolation are of importance.

**Methods:** We assessed nasopharyngeal swabs of COVID-19 patients for quantitative infectious viral titres (IVT) by focus-forming assay and compared to overall virus isolation success and RNA genome copies. We assessed IVTs during the first 5 symptomatic days in a total of 384 patients: unvaccinated individuals infected with pre-VOC SARS-CoV-2 (n= 118) or Delta (n= 127) and vaccine breakthrough infections with Delta (n= 121) or Omicron (n=18).

**Findings:** Correlation between RNA copy number and IVT was low for all groups. No correlation between IVTs and age or sex was seen. We observed higher RNA genome copies in pre-VOC SARS-CoV-2 compared to Delta, but significantly higher IVTs in Delta infected individuals. Vaccinated Delta infected individuals had significantly lower RNA genome copies and IVTs compared to unvaccinated subjects and cleared virus faster. In addition, vaccinated individuals with Omicron infection had comparable IVTs to Delta breakthrough infections.

**Interpretation:** Quantitative IVTs can give detailed insights into virus shedding kinetics. Vaccination was associated with lower infectious titres and faster clearance for Delta, showing that vaccination would also lower transmission risk. Omicron vaccine-breakthrough infections did not show elevated IVTs compared to Delta, suggesting that other mechanisms than increase VL contribute to the high infectiousness of Omicron.

**Funding:** This work was supported by the Swiss National Science Foundation 196644, 196383, NRP (National Research Program) 78 Covid-19 Grant 198412, the Fondation Ancrage Bienfaisance du Groupe Pictet and the Fondation Privée des Hôpitaux Universitaires de Genève.

## Introduction

By 2 January 2021, the coronavirus disease 2019 (COVID-19) pandemic caused nearly 289 million cases and just over 5·4 million deaths globally (1). Severe acute respiratory coronavirus 2 (SARS-CoV-2), the causative agent of COVID-19, primarily infects the cells of the upper respiratory tract (URT) where viral load (VL) increases during the course of infection (2).

The two key measurements of VL are RNA levels, often expressed in cycle threshold (Ct) values, and infectious virus that are assessed by virus isolation in cell culture. Although the transmission process is complex, higher VL can serve as a proxy for greater risk of transmission. In several epidemiological studies, higher VL measured by viral RNA was associated with increased secondary transmission in household settings (3, 4). Infectious SARS-CoV-2 is shed in the URT and starts on average from two days before symptom onset. Even though viral RNA could be detected afterwards, in most studies infectious virus was not detected in respiratory samples collected from immunocompetent individuals later than 8 days post onset of symptoms (DPOS) (5-7). Moreover, viral RNA detection did not correlate with infectiousness in an animal model (8). Instead, isolation success in cell culture was found to correlate with detection of infectious virus from respiratory specimens and the ability to shed and transmit fully competent viral particles (9). Virus isolation success can only give information about the presence or absence of infectious virus, but is not able to quantify the infectious viral titre in samples of the URT (10).

Since the start of the pandemic, SARS-CoV-2 is constantly evolving, leading to the emergence of new variants. While most variants vanished quickly, others such as D614G, and the *variants of concern* (VOCs) Alpha, Beta, Gamma, Delta and Omicron harbour an apparent selection advantage and outcompete other variants locally or even globally. These VOCs exhibit various mutations (11) that lead to immune evasion and/or higher transmissibility, which increased viral shedding (among other factors like environmental stability) can significantly contribute (12, 13). For Alpha, an approximately 10-fold higher RNA VL was observed compared to pre-VOC viral strains, which was correlated with increased isolation success (14, 15). Similarly, Delta also showed 10- to 15-fold higher RNA levels compared to pre-VOC strains (15, 16). However, little is known about the quantity of shed infectious viral particles for VOCs including Omicron.

There is extensive evidence that use of vaccines against SARS-CoV-2, which target the original strain, reduced cases numbers and severity. However, the effect of vaccination on infectious viral shedding and transmission from vaccinated patients remains controversial. All currently approved vaccines are administered intramuscularly, thus the titre of neutralizing antibodies on the mucosal surfaces lining the URT might be limited. Therefore, any sterilizing immunity would probably only be transient (17). Epidemiological studies on the secondary attack rate in households of vaccinated vs unvaccinated index patients led to contradictory results. While some studies show reduced transmission rates from vaccinated index cases (18, 19), one study found no influence of index cases vaccination status (20). However, many additional factors can influence the secondary attack rate in these studies, such as: patient behaviour, age, comorbidities, the infecting variant, time since vaccination and the vaccine used. Therefore, differentiating the effect of vaccination on VL from other factors in purely epidemiological studies is difficult. An overall reduction of RNA VL was reported in vaccinated COVID-19 patients (BNT162b2 mRNA vaccine or ChAdOx1 nCoV-19 (AZD1222) adenoviral vector vaccine), but no difference was observed at 6 months post vaccination (21, 22). Another study found reduced RNA VL early after complete vaccination when Alpha dominated, but no difference at later time points when Delta dominated (23). Additionally, a study investigating the kinetics of RNA VL in COVID-19 patients did not find a difference during the first 5 DPOS, i.e. when most human-to-human transmissions occur, but showed RNA VL declined faster in vaccinated patients (24). Similarly, a lower virus isolation success rate was found in vaccinated vs unvaccinated COVID-19 patients at the same RNA VL, indicating that vaccines can reduce the infectious VL (25). However, no study quantified infectious virus titres of different VOCs in URT samples of vaccinated and unvaccinated COVID-19 patients.

The dynamics of infectious viral shedding in vaccinated and unvaccinated patients infected with relevant VOCs require detailed investigation. Understanding of viral shedding in patients would help shape public health decisions to limit community transmission (26). Here we compare RNA and infectious VL between pre-VOC strains and Delta in unvaccinated patients as well as in vaccination breakthrough infections due to Delta and Omicron. Respiratory samples from mildly symptomatic patients of different age and sex, sampled in the first five DPOS were used for this study. By quantifying infectious viral titres from URT specimens, we show that patients infected with Delta harbour elevated levels of infectious viral titres, while vaccination leads to a reduction of infectious virus.

## Methods

### Participants

#### Sample collection and setting

Nasopharyngeal swabs (NPS) collected from symptomatic individuals in the outpatient testing centre of the Geneva University Hospital, for SARS-CoV-2 RT-PCR diagnostics, were included in this study. Infection with SARS-CoV-2 was diagnosed by RT-PCR assay (Cobas 6800, Roche). All samples originate from the diagnostic unit of the hospital’s virology laboratory and were received for primary diagnosis of SARS-CoV-2. Remaining samples were stored at -80°C, on the same day or within 24h. All samples had only one freeze-thaw cycle for the purpose of this study. All specimens from vaccinated individuals were characterized by full genome sequencing for their infecting SARS-CoV-2 variant. Initial identification of Omicron was done by S-gene target failure of the TaqPath COVID19 assay (Thermofisher) and confirmed by partial Sanger sequencing of Spike (27) followed by next-generation sequencing. No sequence information was obtained for samples collected before the first detection of VOCs in Switzerland, i.e. pre-VOC samples. Clinical information of the patients was collected by a standardized questionnaire in our testing Centre and/or through the Cantonal Health Service.

### Viral load quantification by qRT-PCR

VL in each sample was determined by quantitative real time PCR (RT-qPCR) using SuperScript™ III Platinum™ One-Step qRT-PCR Kit (Invitrogen). RT-PCR for SARS-CoV-2 E gene and quantification of genome copy number was performed as described previously (28).

### Quantification of SARS-CoV-2 by focus-forming assay

Vero E6 and Vero E6-TMPRSS were cultured in complete DMEM GlutaMax I medium supplemented with 10% fetal bovine serum, 1x Non-essential Amino Acids, and 1% antibiotics (Penicillin/Streptomycin) (all reagents from Gibco, USA). Vero-TMPRSS were kindly received from National Institute for Biological Standards and Controls (NIBSC, Cat. Nr. 100978).

NPS samples were serially diluted and applied on a monolayer of VeroE6 cells in duplicates. Following 1 hour at 37°C, the media was removed and prewarmed medium mixed with 2·4% Avicel (DuPont) at a 1:1 ratio was overlaid. Plates were incubated at 37°C for 24 hours and then fixed using 6% paraformaldehyde for 1 hour at room temperature. Cells were permeabilized with 0·1% Triton X-100 and blocked with 1% BSA (Sigma). Plates were incubated with a primary monoclonal antibody targeting SARS-CoV-2 nucleocapsid protein (Geneva Antibody facility; JS02) for 1 hour at room temperature and then with peroxidase-conjugated secondary antibody (Jackson ImmunoResearch, #109-036-09) for 30 minutes at room temperature. Foci were visualized using True Blue HRP substrate (Avantor) and imaged on an ELISPOT reader (CTL). Focus-forming assays for comparison of infectious VLs in Delta vs Omicron were performed in Vero E6-TMPRSS cells.

### Virus isolation

Nasopharyngeal samples were applied on Vero E6 cell monolayers in 24 well plates. 100 µl of each sample was added and incubated for 1 hour at 37°C. Following the incubation, the infectious supernatant was discarded and virus culture medium was added. 50 µL of the medium was collected to determine the VL at day 0. 3-4 days post inoculation the medium was replaced, and 6 days post infection the infectious medium was collected to determine VL. A genome copy number change of at least 1 log of from day 0 to 6 indicated a successful isolation.

### Statistical analysis

All statistical analyses were performed using R Statistical Software version 4.1.1 (Foundation for Statistical 185 Computing, Austria) and Prism version 8.0.1 (GraphPad, San Diego, CA, USA).

### Ethical approval

The study was approved by the Cantonal ethics committee (CCER Nr. 2021-01488). All study participants and/or their legal guardians provided informed consent.

## Results

In this study, we analysed the VL characteristics in the URT of unvaccinated pre-VOC-as well as vaccinated and unvaccinated Delta-infected COVID-19 subjects up to 5 DPOS. We included a total of 384 samples in our cohort of which 118 originated from patients infected with pre-VOC SARS-CoV-2 and 248 from patients infected with the Delta VOC. Of the Delta VOC infected patients, 121 were vaccinated twice prior infection and 127 were unvaccinated. In addition, we included 18 vaccinated individuals recently infected with Omicron. None of the patients infected with pre-VOC SARS-CoV-2 were vaccinated as vaccines were unavailable at the time of infection. All patients had mild symptoms at the time of sampling. Samples of pre-VOC infected patients were collected between April 7^th^ and September 9^th^ 2020, before circulation of any VOCs, samples of Delta-infected patients were collected from June 26^th^ until December 4^th^ 2021, and samples of Omicron-infected patients from 14-17 December 2021. All vaccinated patients included in this study were diagnosed positive at least 14 days after dose 2, which complies with the vaccination breakthrough definition of the Centers for Disease Control and Prevention (29). 132/139 patients were vaccinated with mRNA vaccines, one was vaccinated with a non-replicating viral vector vaccine (CoviVac) and for six patients the vaccine used isn’t known. The median time between 2^nd^ dose and breakthrough infection was 79·5 (IQR 40·5-139 days) for Delta infections and 136 (IQR 85-176) for Omicron infections. All three groups of patients (pre-VOC, Delta-unvaccinated and Delta-vaccinated) had a similar age and sex distribution (see **Table)**.

**Table 1.** Patient characteristics of the specimens used in this study. RT-PCR, reverse transcription polymerase chain reaction, CT, cycle threshold, IQR, interquartile range, na, not applicable. *Of the all 121 vaccine-breakthrough samples, 104 were titrated in parallel with Delta VOC infection and 17 in parallel with Omicron vaccine-break through infections.

|  | Pre-VOC SARS-CoV-2 | SARS-CoV-2 Delta VOC | Vaccine breakthrough infections (Delta VOC) | Vaccine-breakthrough infections (Omicron VOC) |
| --- | --- | --- | --- | --- |
| Number | 118 | 127 | 121* | 18 |
| Sampling dates | April 7 - September 9 2020 | June 26 –August 29 2021 | July 8 -December 4 2021 | December 14 – December 17 |
| Age |  |  |  |  |
| Median (range) | 36 (17-82) | 37 (16-83) | 40 (16-83) | 35 (14-58) |
| <25 | 22 (18.6%) | 19 (14.9 %) | 14 (11.6%) |  |
| 25-35 | 37 (31.4%) | 38 (29.9%) | 36 (29.8%) |  |
| 35-50 | 30 (25.4%) | 41 (32.3%) | 44 (36.3%) |  |
| 50-65 | 23 (19.5%) | 25 (19.7%) | 24 (19.8%) |  |
| >65 | 6 (5.1%) | 4 (3.1%) | 3 (2.5%) |  |
| Sex |  |  |  |  |
| Female | 50 (42.4%) | 65 (51.2%) | 62 (51.2%) | 9 (50 %) |
| Male | 68 (57.6%) | 62 (48.8%) | 59 (48.8%) | 9 (50 %) |
| RT-PCR result, CT (E-gene target, Cobas 6800, Roche) | 13.9-26.6 | 13.8-26.3 | 16.3-26.1 | 17.2-25.9 |
| Interval vaccination to infection, days, mean (IQR) | na | na | 79.5 (IQR 40.5-139 days) | 136 (IQR 85-176) |
| Vaccine |  |  |  |  |
| BNT162b2 | na | na | 43 | 8 |
| mRNA-1273 | na | na | 73 | 8 |
| CoviVac | na | na | 1 | - |
| Vaccine unknown | na | na | 4 | 2 |

We quantified genome copies and infectious viral titres in SARS-CoV-2-positive NPS using qRT-PCR and focus-forming assays. Only specimens with CT-values below 27 for the E-gene RT-PCR diagnostic target (Cobas, Roche), as determined by the clinical laboratory, were included in our study, as previous we and others have shown that infectious virus cannot be reliably isolated from samples with higher CT-values (9, 30). To validate our focus forming assay, we compared it to the ability to successfully isolate virus in cell culture. Virus isolation success has been used as a correlate of infectious viral shedding for SARS-CoV-2 (6, 31-33), but lacks the ability to differentiate between high and low VL samples. We were able to quantify viral titres using the focus forming assay in 91·9%, 91·7% and 83·8% of culture positive samples in the pre-VOC, Delta-unvaccinated, and Delta-vaccinated groups, respectively, indicating a high sensitivity **(Supplementary figure S1A)**. Overall, the Cohens kappa agreement was 0·69, 0·71 and 0·53 for the 3 groups, showing a moderate to substantial agreement **(Supplementary figure S1B)**.

### Low correlation between genome copies and infectious viral titres

First, we investigated whether RNA genome copies are a good proxy for infectious virus shedding. We observed only a very low correlation (R^2^ = 0·119, p=0·0001) between viral genome copies and infectious virus particles for pre-VOC samples **(Figure 1 A)**, while the samples from unvaccinated and vaccinated Delta patients showed slightly higher, yet still low correlations (R^2^ = 0·312, p <0·0001 and R^2^ = 0·399, p <0·0001, respectively) **(Figure 1B, C)**.

**Figure 1.**
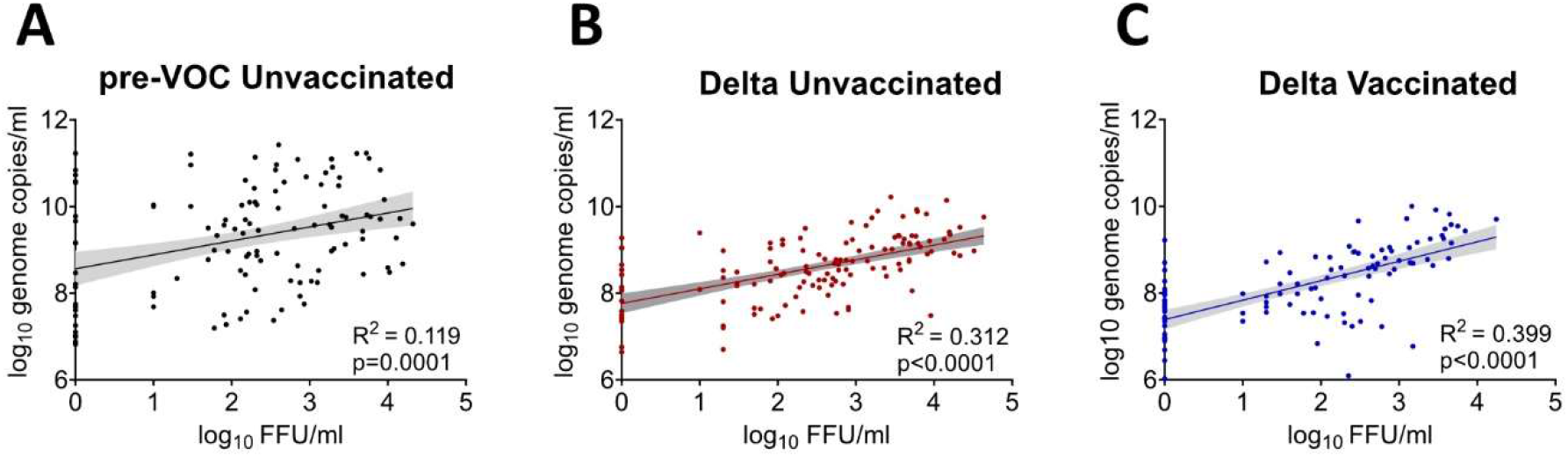
Relationship between RNA viral loads and infectious viral titers. Linear regression analysis of infectious viral titers in FFU/ml and the corresponding RNA viral loads in nasopharyngeal swabs from the unvaccinated patients infected with pre-VOC (**A**), and Delta SARS-CoV-2(**B**) as well as Delta vaccination breakthroughs (**C**).

### No correlation between infectious VL and age and sex of patients

Next, we tested if infectious VLs from patient samples are associated with patient age and sex. We did not observe any correlation between the age and infectious VL for all three groups **(Supplementary figure S2)**. Similarly, no significant differences of infectious VLs between male and female patients were detected for pre-VOC or Delta samples (vaccinated or unvaccinated) **(Supplementary figure S3)**.

### Delta-infected unvaccinated patients have higher infectious VL

Next, we compared genome copies and infectious VLs in pre-VOC and Delta samples from unvaccinated patients during the first 5 DPOS. Overall, pre-VOC samples had significantly more genome copies (4·5 fold, 0·653 log_10_, p<0·0001) compared to Delta samples, but infectious viral titres were significantly higher in Delta-infected individuals (2·2 fold, 0·343 log_10_, p=0·0373) **(Figure 2A)**. We found that genome copies for pre-VOC samples were higher at one and two DPOS, but similar to Delta samples at 0, 3, 4, 5 dpos **(Figure 2B)**. Conversely, infectious virus shedding was higher for Delta at 3-5 DPOS, but similar at 0-2 DPOS **(Figure 2C)**. In addition, we observed that genome copies remained largely stable until 5 DPOS, with only a minimal decline at day 5, while infectious VL substantially declined **(Figure 2B and C)**.

**Figure 2.**
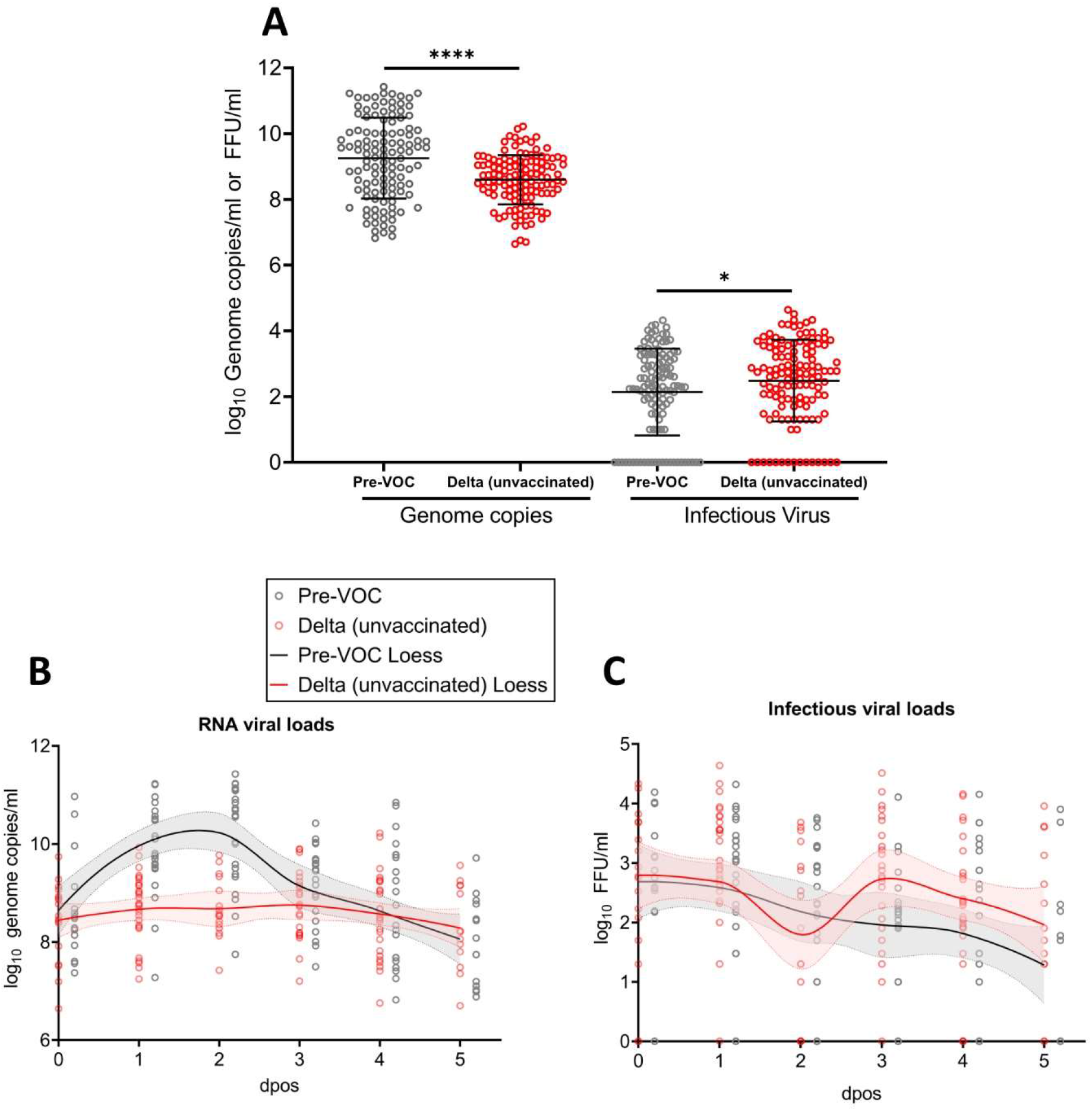
RNA viral load and infectious viral titers for unvaccinated individuals infected with pre-VOC SARS-CoV-2 vs. Delta. **(A)** Genome copies (left panel) and infectious virus (right panel) for pre-VOC and Delta unvaccinated patients. Error bars indicate mean±SD. The t-test was used to determined differences of means. *p=0·0373; **** p<0·0001. Genome copies **(B)** and infectious viral loads **(C)** measured for pre-VOC and Delta VOC infected patients at different dpos. The solid lines represent the fitted curve calculated using (locally estimated scatterplot smoothing) LOESS method.

The association of the infectious shedding levels with patient age and sex is highly debated (14). In this study we also did not detect a correlation between patient age or sex and infectiousness. However, there is increasing evidences of more severe outcomes of COVID-19 disease in older male patients (31, 33). Thus, to eliminate possible confounders, 84 Delta-infected patients were matched with pre-VOC infected patients in regard to sex, age and DPOS. Similarly, significantly higher infectious VLs (3·23 fold, 0·51 log_10_, p=0·001170) were detected in Delta samples compared to matched pre-VOC samples **(Supplementary figure 4A)**.

### Vaccinated patients have lower infectious viral load than unvaccinated patients

To determine vaccination’s influence on virus shedding, we compared genome copies and infectious VLs in unvaccinated and vaccinated patients infected with Delta for 5 DPOS. Overall, RNA genome copies were significantly lower in vaccinated vs. unvaccinated patients (2·5 fold, 0·40 log_10_, p=0·0005). The decrease in infectious VL was even more pronounced in vaccinated patients (4·78 fold, 0·68 log_10_, ****p<0·0001) **(Figure 3A)**. The kinetics of RNA genome copies were largely similar between vaccinated and unvaccinated patients until 3 DPOS with a faster decline for vaccinated patients starting at 4 DPOS **(Figure 3B)**. In contrast, infectious VL were substantially lower in vaccinated patients at all DPOS with the biggest effect at 3-5 DPOS **(Figure 3C)**. Still, at 5 DPOS infectious virus was detectable in 7/13 (53·8%) vaccinated and 11/13 (84·6%) unvaccinated patients. Additionally, 67 Delta-infected patients were matched with Delta vaccine-breakthrough patients in regard to age, sex and dpos. Infectious viral titres were elevated in unvaccinated patients in comparison to vaccine-breakthroughs (9·33 fold, 0·97 log_10_, p<0·0001) **(Supplementary figure S4B)** confirming a significant reduction of infectious VLs among vaccinated patients. We further analysed if there is a correlation between infectious VLs and the time interval since the administration of the last vaccine dose. A high heterogeneity between patient samples resulted in no significant correlation between the time post vaccination and infectious viral shedding **(Supplementary figure S5)**.

**Figure 3.**
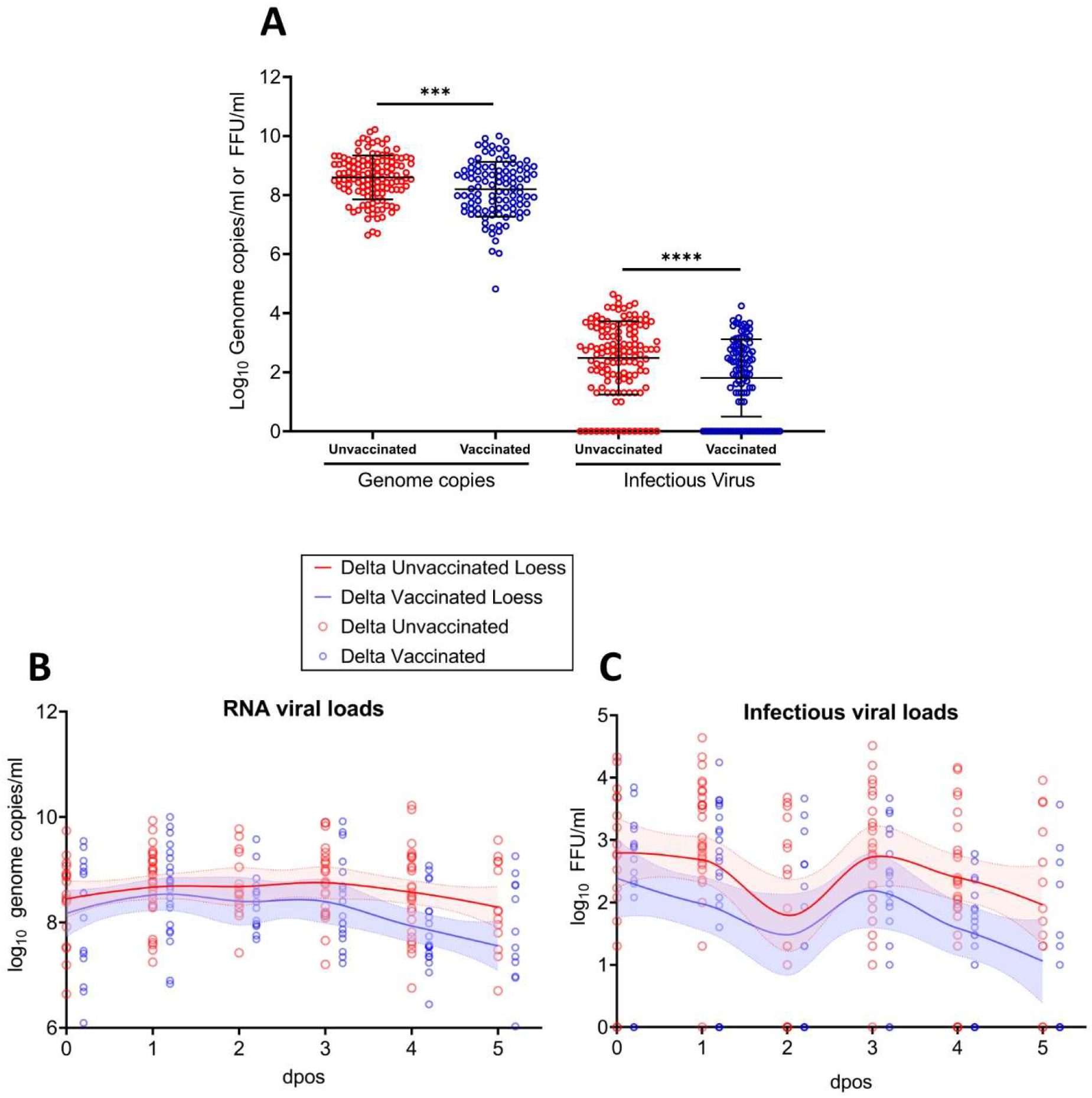
RNA viral load and infectious viral titers for unvaccinated vs. vaccinated individuals infected Delta. **(A)** Genome copies (left panel) and infectious virus (right panel) for vaccinated and unvaccinated Delta infected patients. Error bars indicate mean±SD. The t-test was used to determined differences of means. ***p=0·0005; ****p<0·0001. Genome copies **(B)** and infectious viral loads (C) measured for vaccinated and unvaccinated Delta infected patients at different dpos. The solid lines represent the fitted curve calculated using (locally estimated scatterplot smoothing) LOESS method.

### In previously vaccinated subjects infection with Omicron VOC results in similar infectious viral loads like Delta

Upon the emergence of Omicron, we analysed the infectious viral shedding in vaccinated patients infected with this variant. We compared RNA and infectious VLs in NPS samples of 18 Omicron- and 17 Delta-infected patients. Vaccine-breakthrough infection with Omicron or Delta resulted in comparable genome copies (p= 0·3345). Modestly lower infectious VLs were detected in Omicron-infected patients compared to Delta-infected patients, however this was not statistically significant (4·9 fold, 0·69 log_10_, p= 0·1033) **(Figure 4)**. Similar non-significant reductions of infectious VLs were observed for Omicron samples when matching patients for age, sex and DPOS (**Supplementary figure S4C**).

**Figure 4.**
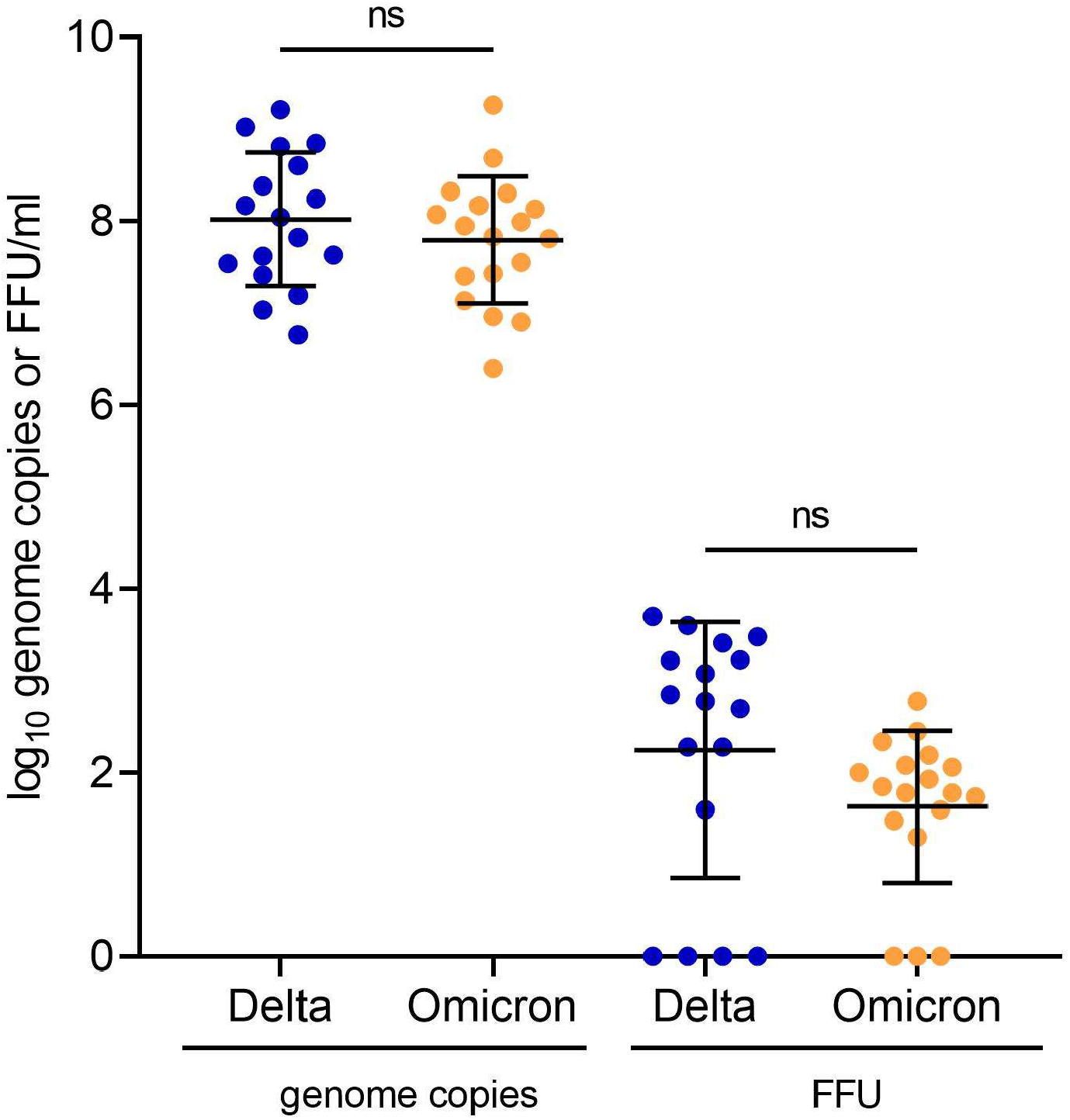
SARS-CoV-2 infectious viral loads in vaccine-break through infections with Omicron or Delta. **(A)** Genome copies (left panel) and infectious virus (right panel) for vaccinated patients infected with Delta or Omicron VOC Infectious viral loads (were determined by focus-forming assay on Vero-TMPRSS cells. Significance was determined by t-test. ns: nonsignificant

## Discussion

In this study we analysed virus shedding in COVID-19 patients infected with pre-VOC, Delta and Omicron variants and evaluated the impact of vaccination on VL in the URT during the first 5 DPOS. To our knowledge, this is the first study which quantified infectious VLs in patients infected with different SARS-CoV-2 variants and vaccination-breakthrough cases. We could demonstrate a higher infectious VL in unvaccinated Delta-infected compared to pre-VOC-infected patients and showed a significant reduction of infectious VLs in vaccinated patients. Furthermore, we found no difference in infectious VL between Delta and Omicron breakthrough cases.

The magnitude and timing of infectiousness of COVID-19 patients is a key requirement to make informed public health decisions on the duration of isolation of patients and on the need to quarantine contacts. Infectiousness is strongly influenced by VL in the URT of infected patients (4). However, VL is often measured as RNA copy number and not actual infectious virus. In this study we could show that RNA copy numbers in NPS samples poorly correlated with infectious virus shedding. This is in line with several other studies that found RNA is a poor infectiousness indicator especially in the presence of neutralising antibodies (9, 32). In addition, in an animal model it was demonstrated that infectious virus, but not RNA, is a good proxy for transmission (8).

Virus isolation in cell culture is widely used as a proxy for infectiousness (6, 9, 28). Several studies have shown that isolation success significantly drops when RNA VLs are below 6 log10 copies per mL in viral transport medium, or samples were collected after 8 DPOS. Of note, with only a qualitative result of successful isolation or not, isolation success cannot distinguish between high and low infectious VLs in a patient sample, a key determinant of the size of the transmitted inoculum. Differences in infectious VL can impact transmission probability, therefore, we used a focus forming assay that can reliably quantify infectious viral particles from patient specimens. Focus forming assays have long been a standard to quantify viral shedding in animal infection models for respiratory viruses such as SARS-CoV-2 and influenza, and are therefore considered one of the best available proxies for infectiousness (34).

Within the first 5 DPOS, we found higher RNA VL in swabs of unvaccinated patients with pre-VOC infections compared to Delta, but infectious VLs were higher for Delta. These results are in disagreement with other studies that analysed only nucleic acid detection and found 3-10-fold higher RNA copy number in Delta-infected patients compared to pre-VOC (15, 35). However, these studies did not control for DPOS, age or sex. Other studies found either no difference in RNA VL between Delta and pre-VOC swabs (36) or more than 1000-fold higher VL for Delta (37), documenting the difficulty of comparing RNA VLs of virus variants during different phases of the pandemic, especially without additional information such as DPOS. Conversely, in agreement with our results, a higher virus isolation success rate was observed for Delta compared to pre-VOC SARS-CoV-2 or Alpha (38).

Vaccines have been shown to tremendously reduce symptomatic SARS-CoV-2 infections. However, vaccination’s impact on breakthrough case infectiousness is unclear. We showed that infectious VL and RNA VL is reduced in vaccinated Delta patients during the first 5 DPOS. In this time period approximately 50% of transmissions occur for pre-VOC strains (5), indicating that reduced VL could considerably decrease secondary attack rate. Other studies showed no difference in RNA VL between the vaccinated and unvaccinated early after symptom onset (24, 25), but found a lower virus isolation rate (25). Conversely, another study detected up to 10-fold reduced RNA VL in vaccinated patients but only for 60 days after complete vaccination (22). Similarly, two more studies reported decreased RNA VL for vaccine-breakthrough infection with pre-VOC and Alpha SARS-CoV-2 (21), but no effect around 6 months post vaccination when Delta dominated (23). Of note, we were still able to detect infectious viral particles in 53·8% of vaccinated subjects at 5 DPOS, indicating that perhaps isolation should not be shortened to 5 days as recommended by the CDC (39). Whether lower infectious VL translates into lower secondary attack rate remains controversial and depends on other influencing factors, e.g. environmental stability of virus particles. Several studies did find a correlation between VL and secondary attack rate, with VL of the index case being the leading transmission correlate (3, 4). In agreement with these findings, epidemiological studies showed reduced transmission from vaccinated index cases, but the effect size depends on the prevalent variant, the vaccine used and the time since vaccination (18). In contrast, another study found that the index case vaccination status did not influence the secondary attack rate (20). While VL is a key element of transmission, the process of human-to-human transmission is complex and other factors, such as varying recommended protection measures, overall incidence, perceived risks and the context of contacts (household *vs* community transmission) can influence outcomes in the studies reported.

To date, few data exist on VL in vaccine-breakthrough infections caused by Omicron due to its recent emergence in late November 2021. Reduced neutralization of Omicron by infection- and vaccine-derived antibodies was reported *in vitro* and epidemiological studies show an increased risk of (re-) infection with Omicron in vaccinated and recovered individuals (40, 41). Furthermore, very high transmissibility of Omicron breakthrough infections was observed, with high secondary attack rates among vaccinated individuals (42). Higher RNA VLs as described in some studies were discussed as one potential contributing factor for the emergence of Alpha and Delta, although for Delta we could only confirm this for infectious VL in our data. The contribution of VL to the transmissibility of Omicron is currently unknown, nor is the mechanism behind the higher transmissibility. First in vitro data hint towards alternative entry mechanisms as well as early replication peaks in cell culture (43), but no clinical data for these exist so far. Our findings indicate that with comparable RNA VL as well as comparable infectious VL, the higher transmissibility in Omicron seems to be unrelated to an increased shedding of infectious viral particles in vaccinated individuals.

Our study has several limitations. We included only samples collected <5 DPOS with Ct-values <27. Therefore, absolute RNA copy numbers are biased towards higher VLs as patients with low VL were not included here. However, patients with low VL have likely little relevance in terms of transmission. Other factors, such as poor swab quality can be a confounding factor leading to low VLs. Furthermore, our focus was on infectious virus shedding and it has been shown that SARS-CoV-2 culture is unlikely to be successful from samples with higher Ct-values (30) and that the vast majority of secondary transmission occurs before 5 dpos although this requires assessment in Omicron cases (5). Due to its recent emergence, we did not yet have access to samples from Omicron-infected unvaccinated individuals. Lastly, we also would like to emphasize that almost all patients in this study were vaccinated with mRNA vaccines that induce high titres of neutralizing antibodies in the blood but relatively low mucosal antibodies. Therefore, our results cannot be generalized to other vaccines, i.e. those that are used mainly in low- and middle-income countries.

In conclusion, this study provides strong evidence for higher infectiousness of the Delta as well as a significantly lower infectiousness and a faster clearance of infectious virus in vaccinated individuals. In addition, we could show that Omicron has similar infectious VLs to Delta. Furthermore, we show a more detailed picture of VL assessment in addition to overall isolation success, and that quantifying VLs can give better insights into shedding kinetics in acute SARS-CoV-2 infections.

## Data Availability

All data produced in the present study are available upon reasonable request to the authors

## Acknowledgments

We thank all patients for their willingness to participate in our research. We thank the staff of the laboratory of virology from the University Hospitals of Geneva for their support. We also thank Eric Boehm for help with editing the manuscript.

## Data sharing

Anonymized data can be made available upon reasonable request.

## Supplementary figures

**Supplementary figure S1.**
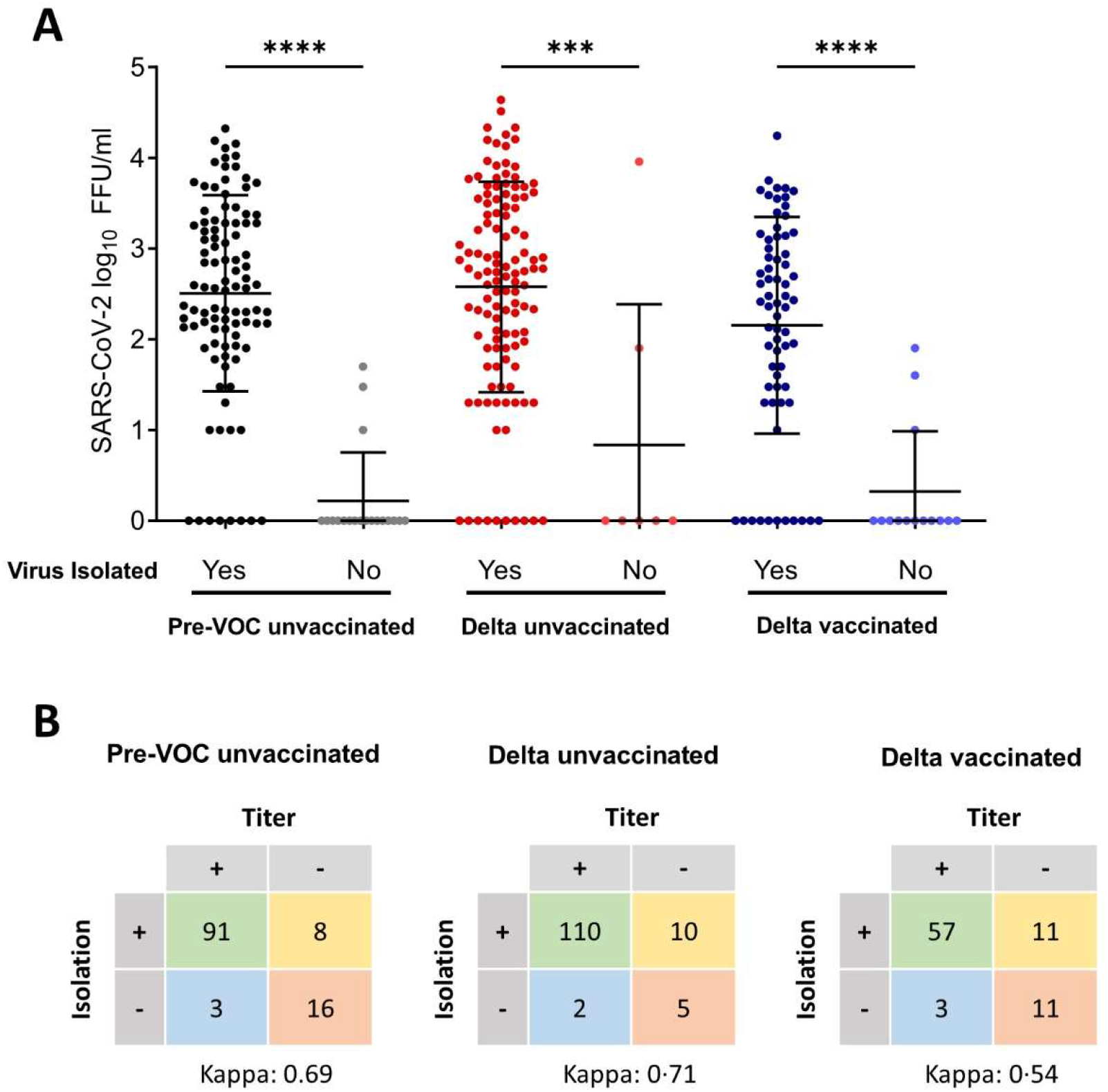
Quantitative infectious viral loads versus overall virus isolation success. (**A**) Vero E6 cells were inoculated with 10-fold serial dilutions of nasopharyngeal swabs collected from SARS-CoV-2 infected individuals. Plates were fixed 27 h post-infection and following the staining with SARS-CoV-2 specific antibodies, the number of focus forming units (FFU)/ml was calculated for each sample. Error bars indicate mean±SD. p-values were calculated with the one-way ANOVA. ***: p<0·002; ****p<0·0001. (**B**) The total number of positive and negative samples defined by titration and virus isolation for each patient group. Cohens kappa agreement is shown.

**Supplementary figure S2.**
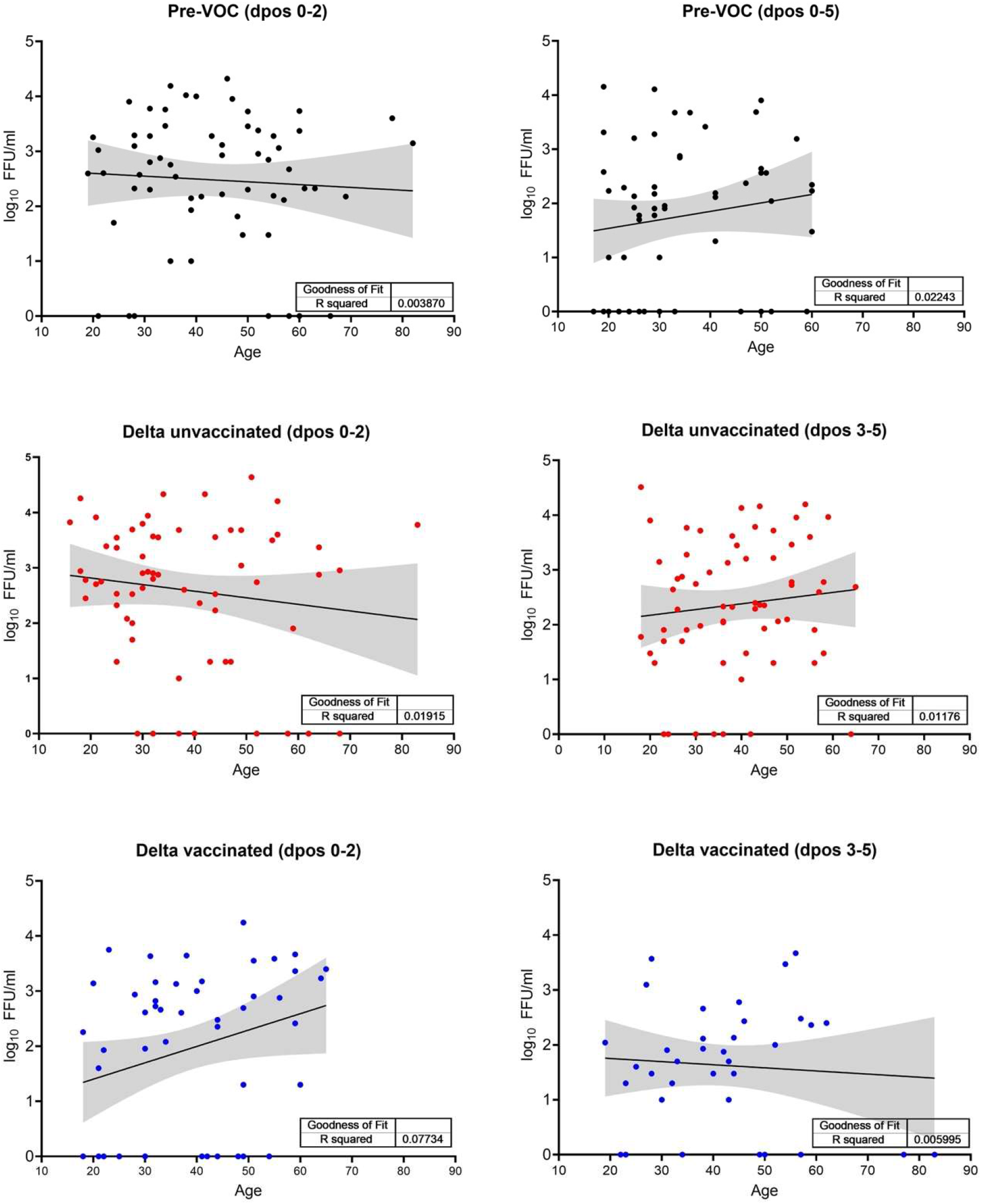
Linear regression analysis of SARS-CoV-2 titers in FFU/ml and the corresponding age of the patient.

**Supplementary figure S3.**
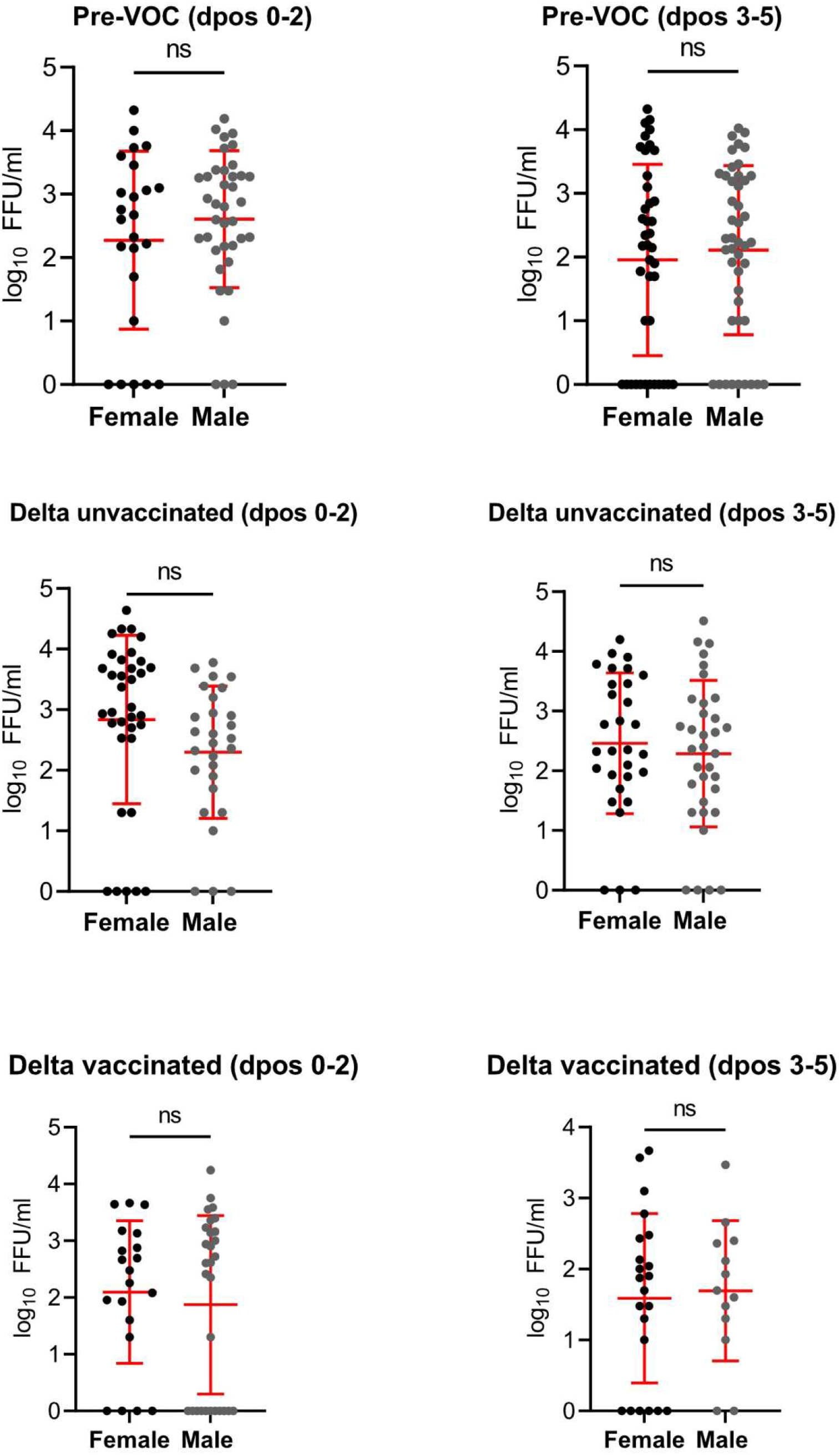
Comparison of infectious viral shedding measured in female and male patients. Error bars indicate mean±SD. The t-test was used to determine differences of means. ns= nonsignificant.

**Supplementary figure S4.**
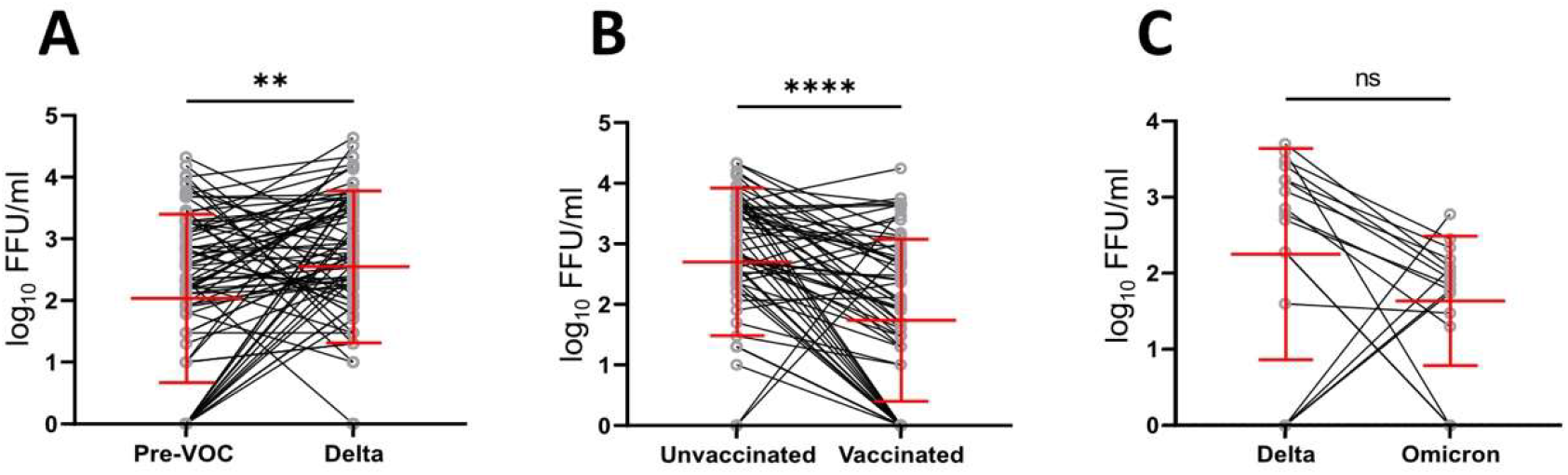
SARS-CoV-2 infectious viral loads detected in unvaccinated patients infected with pre-VOC or Delta (A), unvaccinated and vaccinated patients infected with Delta (B), vaccinated patients infected with Delta or Omicron (C) matched by age, sex, and dpos. Error bars indicate mean±SD. **p=0·001170; ****p<0·0001, ns=nonsignificant.

**Supplementary Figure S5.**
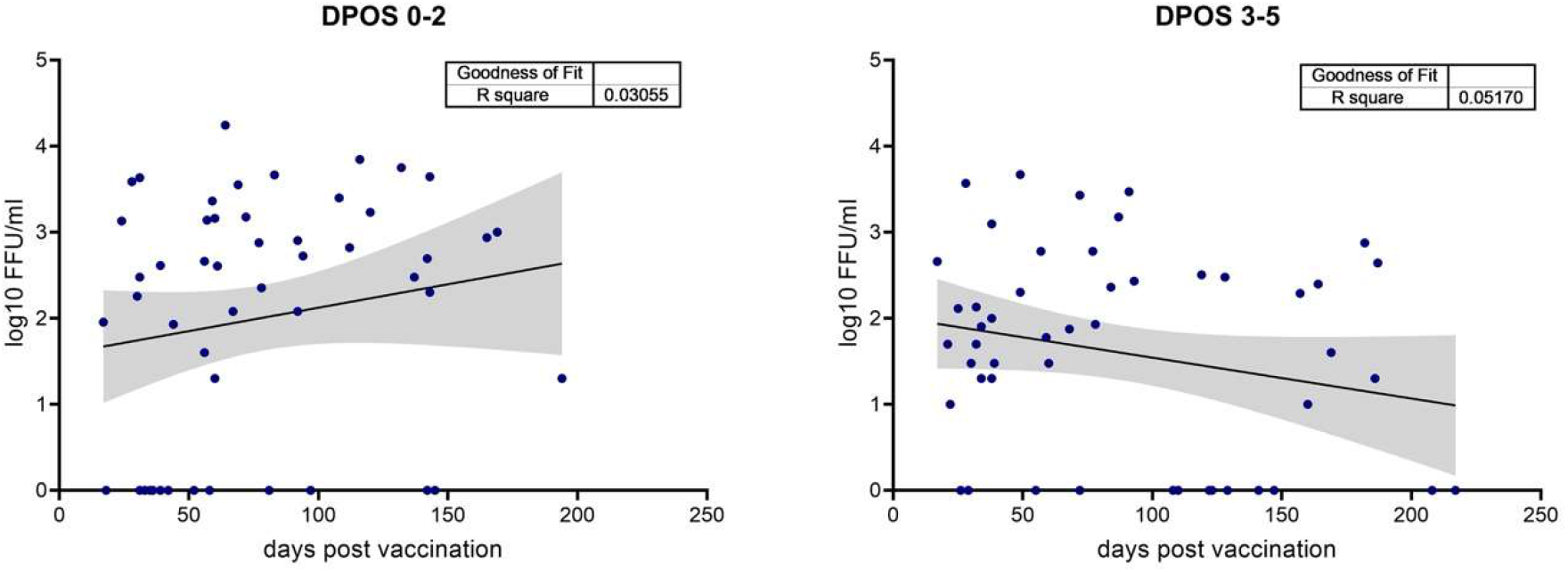
Linear regression analysis of infectious viral shedding and time since the completion of vaccination in Delta infected patients.

## References

1. Weekly epidemiological update on COVID-19 - 6 January 2022. World Health Organization; 2022.

2. Kawasuji H, Takegoshi Y, Kaneda M, Ueno A, Miyajima Y, Kawago K, et al. Transmissibility of COVID-19 depends on the viral load around onset in adult and symptomatic patients. PLoS One. 2020;15(12):e0243597.

3. Marc A, Kerioui M, Blanquart F, Bertrand J, Mitjà O, Corbacho-Monné M, et al. Quantifying the relationship between SARS-CoV-2 viral load and infectiousness. Elife. 2021;10.

4. Marks M, Millat-Martinez P, Ouchi D, Roberts CH, Alemany A, Corbacho-Monné M, et al. Transmission of COVID-19 in 282 clusters in Catalonia, Spain: a cohort study. Lancet Infect Dis. 2021;21(5):629–36.

5. He X, Lau EHY, Wu P, Deng X, Wang J, Hao X, et al. Temporal dynamics in viral shedding and transmissibility of COVID-19. Nat Med. 2020;26(5):672–5.

6. Wölfel R, Corman VM, Guggemos W, Seilmaier M, Zange S, Müller MA, et al. Virological assessment of hospitalized patients with COVID-2019. Nature. 2020;581(7809):465–9.

7. Vetter P, Eberhardt CS, Meyer B, Martinez Murillo PA, Torriani G, Pigny F, et al. Daily Viral Kinetics and Innate and Adaptive Immune Response Assessment in COVID-19: a Case Series. mSphere. 2020;5(6).

8. Sia SF, Yan L-M, Chin AWH, Fung K, Choy K-T, Wong AYL, et al. Pathogenesis and transmission of SARS-CoV-2 in golden hamsters. Nature. 2020;583(7818):834–8.

9. van Kampen JJA, van de Vijver DAMC, Fraaij PLA, Haagmans BL, Lamers MM, Okba N, et al. Duration and key determinants of infectious virus shedding in hospitalized patients with coronavirus disease-2019 (COVID-19). Nature Communications. 2021;12(1):267.

10. Despres HW, Mills MG, Shirley DJ, Schmidt MM, Huang M-L, Jerome KR, et al. Quantitative measurement of infectious virus in SARS-CoV-2 Alpha, Delta and Epsilon variants reveals higher infectivity (viral titer:RNA ratio) in clinical samples containing the Delta and Epsilon variants. medRxiv. 2021:2021.09.07.21263229.

11. Rapid risk assessment: Assessing SARS-CoV-2 circulation, variants of concern, non-pharmaceutical interventions and vaccine rollout in the EU/EEA, 15th update. European Centre for Disease Prevention and Control; 2021.

12. Xia S, Wen Z, Wang L, Lan Q, Jiao F, Tai L, et al. Structure-based evidence for the enhanced transmissibility of the dominant SARS-CoV-2 B.1.1.7 variant (Alpha). Cell Discov. 2021;7(1):109.

13. Dejnirattisai W, Zhou D, Supasa P, Liu C, Mentzer AJ, Ginn HM, et al. Antibody evasion by the P.1 strain of SARS-CoV-2. Cell. 2021;184(11):2939-54.e9.

14. Jones TC, Biele G, Mühlemann B, Veith T, Schneider J, Beheim-Schwarzbach J, et al. Estimating infectiousness throughout SARS-CoV-2 infection course. Science. 2021;373(6551).

15. Teyssou E, Delagrèverie H, Visseaux B, Lambert-Niclot S, Brichler S, Ferre V, et al. The Delta SARS-CoV-2 variant has a higher viral load than the Beta and the historical variants in nasopharyngeal samples from newly diagnosed COVID-19 patients. J Infect. 2021;83(4):e1–e3.

16. Imai K, Ikeno R, Tanaka H, Takada N. SARS-CoV-2 Delta variant saliva viral load is 15-fold higher than wild-type strains. medRxiv. 2021:2021.11.29.21266980.

17. Mostaghimi D, Valdez CN, Larson HT, Kalinich CC, Iwasaki A. Prevention of host-to-host transmission by SARS-CoV-2 vaccines. Lancet Infect Dis. 2021.

18. Harris RJ, Hall JA, Zaidi A, Andrews NJ, Dunbar JK, Dabrera G. Effect of Vaccination on Household Transmission of SARS-CoV-2 in England. N Engl J Med. 2021;385(8):759–60.

19. Eyre DW, Taylor D, Purver M, Chapman D, Fowler T, Pouwels KB, et al. Effect of Covid-19 Vaccination on Transmission of Alpha and Delta Variants. N Engl J Med. 2022.

20. Singanayagam A, Hakki S, Dunning J, Madon KJ, Crone MA, Koycheva A, et al. Community transmission and viral load kinetics of the SARS-CoV-2 delta (B.1.617.2) variant in vaccinated and unvaccinated individuals in the UK: a prospective, longitudinal, cohort study. Lancet Infect Dis. 2021.

21. Emary KRW, Golubchik T, Aley PK, Ariani CV, Angus B, Bibi S, et al. Efficacy of ChAdOx1 nCoV-19 (AZD1222) vaccine against SARS-CoV-2 variant of concern 202012/01 (B.1.1.7): an exploratory analysis of a randomised controlled trial. Lancet. 2021;397(10282):1351–62.

22. Levine-Tiefenbrun M, Yelin I, Alapi H, Katz R, Herzel E, Kuint J, et al. Viral loads of Delta-variant SARS-CoV-2 breakthrough infections after vaccination and booster with BNT162b2. Nature Medicine. 2021;27(12):2108–10.

23. Pouwels KB, Pritchard E, Matthews PC, Stoesser N, Eyre DW, Vihta K-D, et al. Effect of Delta variant on viral burden and vaccine effectiveness against new SARS-CoV-2 infections in the UK. Nature Medicine. 2021;27(12):2127–35.

24. Chia PY, Ong SWX, Chiew CJ, Ang LW, Chavatte JM, Mak TM, et al. Virological and serological kinetics of SARS-CoV-2 Delta variant vaccine breakthrough infections: a multicentre cohort study. Clin Microbiol Infect. 2021.

25. Shamier MC, Tostmann A, Bogers S, de Wilde J, IJpelaar J, van der Kleij WA, et al. Virological characteristics of SARS-CoV-2 vaccine breakthrough infections in health care workers. medRxiv. 2021:2021.08.20.21262158.

26. Badu K, Oyebola K, Zahouli JZB, Fagbamigbe AF, de Souza DK, Dukhi N, et al. SARS-CoV-2 Viral Shedding and Transmission Dynamics: Implications of WHO COVID-19 Discharge Guidelines. Front Med (Lausanne). 2021;8:648660.

27. Sabine Yerly LK, Manuel Schibler, Isabella Eckerle. Protocol for specific RT-PCRs for marker regions of the Spike indicative of the Omicron variant (B.1.1.529). Geneva, Switzerland: Centre for Emerging Viral Diseases, Geneva University Hospitals; December 2,2021.

28. Corman VM, Landt O, Kaiser M, Molenkamp R, Meijer A, Chu DK, et al. Detection of 2019 novel coronavirus (2019-nCoV) by real-time RT-PCR. Euro Surveill. 2020;25(3).

29. COVID-19 Vaccine Breakthrough Infections Reported to CDC — United States, January 1– April 30, 2021. 2021 May 28. 2021.

30. Essaidi-Laziosi M, Perez Rodriguez FJ, Hulo N, Jacquerioz F, Kaiser L, Eckerle I. Estimating clinical SARS-CoV-2 infectiousness in Vero E6 and primary airway epithelial cells. Lancet Microbe. 2021;2(11):e571.

31. Chen PZ, Bobrovitz N, Premji ZA, Koopmans M, Fisman DN, Gu FX. SARS-CoV-2 shedding dynamics across the respiratory tract, sex, and disease severity for adult and pediatric COVID-19. Elife. 2021;10.

32. Jefferson T, Spencer EA, Brassey J, Heneghan C. Viral Cultures for Coronavirus Disease 2019 Infectivity Assessment: A Systematic Review. Clin Infect Dis. 2021;73(11):e3884–e99.

33. Takahashi T, Ellingson MK, Wong P, Israelow B, Lucas C, Klein J, et al. Sex differences in immune responses that underlie COVID-19 disease outcomes. Nature. 2020;588(7837):315–20.

34. Wong L-YR, Li K, Sun J, Zhuang Z, Zhao J, McCray PB, et al. Sensitization of Non-permissive Laboratory Mice to SARS-CoV-2 with a Replication-Deficient Adenovirus Expressing Human ACE2. STAR Protocols. 2020;1(3):100169.

35. Christian von Wintersdorff JD, Lieke van Alphen, Petra Wolffs, Brian van der Veer, Christian Hoebe, Paul Savelkoul. Infections caused by the Delta variant (B.1.617.2) of SARS-CoV-2 are associated with increased viral loads compared to infections with the Alpha variant (B.1.1.7) or non-Variants of Concern2021.

36. Tani-Sassa C, Iwasaki Y, Ichimura N, Nagano K, Takatsuki Y, Yuasa S, et al. Viral loads and profile of the patients infected with SARS-CoV-2 Delta, Alpha, or R.1 variants in Tokyo. J Med Virol. 2021.

37. Wang Y, Chen R, Hu F, Lan Y, Yang Z, Zhan C, et al. Transmission, viral kinetics and clinical characteristics of the emergent SARS-CoV-2 Delta VOC in Guangzhou, China. EClinicalMedicine. 2021;40:101129.

38. Luo CH, Morris CP, Sachithanandham J, Amadi A, Gaston DC, Li M, et al. Infection with the SARS-CoV-2 Delta Variant is Associated with Higher Recovery of Infectious Virus Compared to the Alpha Variant in both Unvaccinated and Vaccinated Individuals. Clin Infect Dis. 2021.

39. Prevention CfDCa. CDC Updates and Shortens Recommended Isolation and Quarantine Period for General Population. December 27, 2021.

40. Eggink D, Andeweg SP, Vennema H, van Maarseveen N, Vermaas K, Vlaemynck B, et al. Increased risk of infection with SARS-CoV-2 Omicron compared to Delta in vaccinated and previously infected individuals, the Netherlands, 22 November to 19 December 2021. medRxiv. 2021:2021.12.20.21268121.

41. Pulliam JRC, van Schalkwyk C, Govender N, von Gottberg A, Cohen C, Groome MJ, et al. Increased risk of SARS-CoV-2 reinfection associated with emergence of the Omicron variant in South Africa. medRxiv. 2021:2021.11.11.21266068.

42. Brandal LT, MacDonald E, Veneti L, Ravlo T, Lange H, Naseer U, et al. Outbreak caused by the SARS-CoV-2 Omicron variant in Norway, November to December 2021. Euro Surveill. 2021;26(50).

43. Peacock TP, Brown JC, Zhou J, Thakur N, Newman J, Kugathasan R, et al. The SARS-CoV-2 variant, Omicron, shows rapid replication in human primary nasal epithelial cultures and efficiently uses the endosomal route of entry. bioRxiv. 2022:2021.12.31.474653.

